# Temporal tau asymmetry spectrum influences divergent behavior and language patterns in Alzheimer’s disease

**DOI:** 10.1101/2023.11.10.23296836

**Authors:** Kyan Younes, Viktorija Smith, Emily Johns, Mackenzie L. Carlson, Joseph Winer, Zihuai He, Victor W. Henderson, Michael D. Greicius, Christina B. Young, Elizabeth C. Mormino, Alzheimer’s Disease Neuroimaging Initiative Researchers

**Affiliations:** Department of Neurology and Neurological Sciences, Stanford University School of Medicine; Quantitative Sciences Unit, Department of Medicine, Stanford University, Stanford, California; Department of Epidemiology and Population Health, Stanford University; Wu Tsai Neuroscience Institute, Stanford, CA, USA

**Author notes:** Corresponding Author: Kyan Younes, MD, 213 Quarry Road, Palo Alto, CA 94304.

## Abstract

Understanding psychiatric symptoms in Alzheimer’s disease (AD) is crucial for advancing precision medicine and therapeutic strategies. The relationship between AD behavioral symptoms and asymmetry in spatial tau PET patterns is unknown. Braak tau progression implicates the temporal lobes early. However, the clinical and pathological implications of temporal tau laterality remain unexplored.

This cross-sectional study investigated the correlation between temporal tau PET asymmetry and behavior assessed using the neuropsychiatric inventory, and composite scores for memory, executive function, and language; using data from the Alzheimer’s Disease Neuroimaging Initiative (ADNI) dataset. In the entire cohort, continuous right and left temporal tau contributions to behavior and cognition were evaluated controlling for age, sex, education, and tau burden on the contralateral side. Additionally, a temporal tau laterality index was calculated to define “asymmetry-extreme” groups (individuals with laterality indices greater than two standard deviations from the mean).

858 individuals (age=73.9±7.7 years, 434(50%) females) were included, comprising 438 cognitively unimpaired (CU) (53.4%) and 420 impaired (CI) participants (48.9%). In the full cohort analysis, right temporal tau was associated with worse behavior (*B*(SE)=7.19 (2.9), *p-value*=0.01) and left temporal tau was associated with worse language (*B*(SE)=1.4(0.2), *p-value*<0.0001). Categorization into asymmetry-extreme groups revealed 20 right- and 27 left-asymmetric participants. Within these extreme groups, four patterns of tau PET uptake were observed: anterior temporal, typical AD, typical AD with frontal involvement, and posterior.

Asymmetrical tau burden is associated with distinct behavioral and cognitive profiles. Behavioral and socioemotional measures are needed to understand right-sided asymmetry in AD.

## Introduction

Beta-amyloid plaques and tau neurofibrillary tangles are the hallmark pathological findings in Alzheimer’s disease (AD).^1^ Unlike amyloid, the spatial distribution of tau correlates with specific clinical symptoms, domain-specific cognitive impairments, and dementia severity.^2,3^ While the burden and spatial distribution of tau have been extensively studied in relation to cognitive symptoms, the link between tau distribution and behavioral and emotional symptoms remains underexamined. Similarly, while the focus of disease modifying therapies is improving cognitive symptoms, behavioral and emotional endpoints are rarely considered.

Emotional and behavioral changes represent a major challenge to patients,^4^ caregivers,^5^ and the health care system.^6^ Insidious behavioral and emotional changes occur commonly in the mild cognitive impairment (MCI) and dementia stages of AD.^7,8^ MCI is defined as impairment in one or more cognitive domains,^9^ overlooking the early emotional and behavioral changes commonly reported in patients with AD.^7,8^ The stereotypical appearance of tau pathology in sporadic AD involves the transentorhinal region followed by further spread within the medial temporal lobe and subsequently into neocortical regions.^10,11^ However, many neuropathological and neuroimaging studies highlight heterogeneity in the spatial pattern of neocortical tau tangle deposition and have identified asymmetric subtypes.^3,12–18^ While asymmetric tau distribution is known in atypical AD presentations, temporal lobe tau asymmetry in typical amnestic AD has been understudied. Importantly, in addition to memory, the temporal lobes subserve aspects of language and behavior.^19–23^

Although understudied in AD, socioemotional and behavioral symptoms have been deeply characterized in frontotemporal dementia (FTD), which presents with various combinations of behavioral, personality, mood, cognitive, and motor symptoms.^24,25^ Notably, left- or right-predominant anterior temporal atrophy is associated with semantic variant primary progressive aphasia (svPPA) or semantic behavioral variant frontotemporal dementia (sbvFTD), respectively, and on the molecular levels both svPPA and sbvFTD show vulnerability to sporadic TAR DNA-binding protein-43 (TDP-43) type C pathology.^26^ It is well established that left anterior temporal lobe involvement is associated with verbal semantic loss whereas right anterior temporal lobe involvement is associated with non-verbal and socioemotional deficits. ^10,11^ We hypothesized that a similar pattern of graded right-socioemotional/behavioral to left-linguistic spectrum will be present in AD depending on asymmetric tau distribution in the temporal lobes. To address this hypothesis, we leveraged the Alzheimer’s Disease Neuroimaging Initiative (ADNI) study to quantify asymmetric temporal tau burden using tau PET imaging and investigated the behavioral and cognitive patterns associated with right and left predominant tau burden.

### Methods and Materials Participants

We included ADNI participants with available tau PET (n=864; UCBERKELEYAV1451_PVC_04_29_22.csv downloaded at LONI, https://ida.loni.usc.edu; Appendix for information on ADNI). Demographics including age, sex, education, and handedness were extracted from ADNI datasheets. Participants’ first tau PET scan was included when more than one scan was available. All ADNI participants provided written informed consent in compliance with local IRBs.

### Functional and neuropsychosocial measures

The clinical dementia rating (CDR)^27^ scale was obtained through a semi-structured interview of the participant and study partner and provides a measure of functional impairment. The cognitive outcome measures were harmonized composite memory, language, and executive function scores.^28^ The behavioral outcome was the Neuropsychiatric Inventory (NPI) total and subdomain scores.^29^ The NPI probes the severity and frequency of multiple behaviors including delusions, hallucinations, agitation/aggression, dysphoria, anxiety, euphoria, apathy, disinhibition, irritability/lability, aberrant motor activity, night-time behavioral disturbances, appetite, and eating abnormalities.

### Genetic data

Genetic data were downloaded from the ADNI website to investigate common genetic variants in neurodegenerative disease. Available ADNI genetic data for the subjects included in our study was queried for the following genetic mutations: *PGRN, MAPT, TARDBP, C9orf72, APP, PSEN1, PSEN2, FUS* and *APOE*.

### Temporal tau laterality index

We examined temporal tau PET SUVR values accessible on LONI (inferior cerebellum reference region) and Amyloid PET status accessible on LONI (whole cerebellum reference region). Right and left temporal ROIs included entorhinal, fusiform, inferior temporal, middle temporal, superior temporal, temporal pole, transverse temporal, and medial temporal lobe (MTL; comprising hippocampus, parahippocampal, and amygdala). We additionally calculated a left temporal tau SUVR and a right temporal tau SUVR by averaging the SUVRs of all the right and left temporal ROIs and a whole temporal ROI by averaging all the temporal ROIs. To examine the effect of tau asymmetry on cognition and behavior in the entire cohort and the effect of tau asymmetry subgroups, we performed two sets of complementary analyses:

1. <u>Asymmetry-extreme groups</u> analysis of the cases falling greater than two standard deviations away from the mean of tau laterality index (LI) across the entire sample. LI was calculated using the following equation (Left ROI SUVR – Right ROI SUVR)/(Left ROI SUVR + Right ROI SUVR).^21,30,31^ The LI mean and standard deviation of the entire sample were calculated, and participants were considered left-predominant if they were over two standard deviations above the mean and right-predominant if they were two standard deviations below the mean on the LI.
2. <u>Asymmetry-spectrum</u> analysis was performed to investigate the influence of continuous tau laterality across the entire cohort. For this approach, we included both right and left temporal tau SUVRs in the same model, to identify effects of each hemisphere controlling for the contralateral hemisphere (e.g., behavior/cognition ∼ age + sex + education + right temporal tau SUVR + left temporal tau SUVR). This analysis was done using the temporal ROIs as well as using the individual ROIs within the temporal lobe.

### Voxelwise Tau PET

For participants classified as asymmetry-extreme, image data for Tau PET [[18]F-flortaucipir (FTP)] and the structural MRI scan obtained closest in time to Tau PET were downloaded from LONI. Summed Tau PET images were selected with the file description ‘AV1451Coreg, Avg, Std Img and Vox Siz, Uniform 6mm Res’. Tau PET data were processed with in-house scripts using SPM and FSL. Each summed FTP image was co-registered to that participant’s corresponding MRI scan. The MRI scan was spatially normalized with the Tissue Probability map and these transformations were applied to the co-registered PET image. The mean inferior cerebellum was defined by the Normalized Probability Desikan-Killiany Atlas^32^ and used as the reference region to calculate standardized uptake value ratios (SUVR) on the resulting warped images. Conjunction and average maps of the right-predominant group, left-predominant group and the combined right-predominant and left-predominant groups were created using a tau SUVR threshold of 0.25 (i.e., masks for tau SUVR values over 0.25 were created, binarized, and summed to create conjunction maps).

### Statistical analysis

Data analysis was performed with R version 4.0.2. Normality for continuous data was tested with the Shapiro–Wilk test. Means for continuous variables were compared with the Student’s t or the Mann-Whitney U tests. Baseline characteristics were compared using the χ2 or Fisher exact test for categorical variables. Homogeneity of variance was tested by Levene’s test. Given the majority of participants with clinical impairment were in the MCI stage and the number of dementia and MCI-other individuals was low (**Table 1**), we combined all impaired individuals into one cognitively impaired (CI) group. Statistical significance was determined at *p-value*<0.05.

**Table 1.** Demographics, functional, diagnostic, APOE4 and Amyloid status information of the asymmetry-spectrum and asymmetry-extreme groups. HW: Hispanic White; NHW: Non-Hispanic White, NHA: Non-Hispanic Asian, NHB: Non-Hispanic Black, AIAN, UNK, +2 more than 2.

|  | Right index spectrum | Left index spectrum | Total laterality spectrum | Right extreme | Left extreme |
| --- | --- | --- | --- | --- | --- |
| Count | 393 (46) | 465 (54) | 858 | 20 | 27 |
| Age mean (sd) | 73.8 (7.8) | 73.9 (7.7) | 73.9 (7.7) | 74.96 (5.67) | 72.01 (6.16) |
| Handedness n (%) | R = 369 (94)<br>L = 18 (4.5)<br>Amb = 5 (1.3)<br>UKN = 1 (0.2) | R = 434 (93.5)<br>L = 21 (4.5)<br>Amb = 10 (2.2)<br>UKN = 0 | R = 804 (93.7)<br>L = 39 (4.5)<br>Amb = 15 (1.7)<br>UKN = 1 (0.1) | R = 18 (90)<br>L = 2 (10) | R = 22 (82)<br>L = 5 (18) |
| Education mean (sd) | 16.5 (2.5) | 16.3 (2.4) | 16.4 (2.4) | 16.55 (2.74) | 15.88 (2.43) |
| Female/Male | F = 187 / M = 203 | F = 247 / M = 218 | F = 434/ M = 424 | F = 7 / M = 13 | F = 13 / M = 14 |
| Race/ethnicity n (%) | HW: 14 (3.7)<br>NHW: 325 (82.7)<br>NHA: 10 (2.5)<br>NHB: 32 (8.1)<br>AIAN: 1 (0.2)<br>+2: 3 (0.8)<br>UNK: 8 (2) | HW: 25 (5.4)<br>NHW: 385 (82.8)<br>NHA: 12 (2.6)<br>NHB: 28 (6.1)<br>AIAN: 1 (0.2)<br>+2: 7 (1.5)<br>UNK: 7 (1.5) | HW: 39 (4.5)<br>NHW: 710 (82.7)<br>NHA: 22 (2.6)<br>NHB: 60 (7.0)<br>AIAN: 2 (0.2)<br>+2: 2: 10 (1.2)<br>UNK: 15 (1.7) | NHW: 20 (100) | NHW: 27 (100) |
| APOE4 n (%) | 0 = 210 (53.4)<br>1 = 118 (30)<br>2 = 24 (6.1)<br>NA = 41 (10.4) | 0 = 253 (54.4)<br>1 = 129 (27.7)<br>2 = 34 (7.3)<br>NA = 49 (10.5) | 0 = 463 (53.9)<br>1 = 247 (28.8)<br>2 = 58 (6.8)<br>NA = 90 (10.5) | 0 = 6 (30)<br>1 = 8 (40)<br>2 = 3 (15)<br>NA = 3 (15) | 0 = 7 (25.9)<br>1 = 13 (48.1)<br>2 = 7 (25.9) |
| CDR mean (sd) | 0.25 (0.34) | 0.29 (0.41) | 0.27 (0.38) | 3.20 (2.49) | 3.12 (3.34) |
| Diagnosis n (%) | CN = 219 (55.7)<br>MCI-AD = 157 (39.9)<br>MCI-other = 12 (3.1)<br>Dementia = 4 (1)<br>Missing = 1 (0.2) | CN = 239 (51.4)<br>MCI-AD = 199 (42.8)<br>MCI-other = 10 (2.2)<br>Dementia = 6 (1.3)<br>Missing = 11 (2.4) | CN = 458 (53.4)<br>MCI-AD = 356 (41.5)<br>MCI-other = 22 (2.6)<br>Dementia = 10 (1.2)<br>Missing = 12 (1.4) | CN = 3 (15)<br>MCI-AD = 16 (80)<br>MCI-other = 1 (5) | CN = 6 (22.2)<br>MCI-AD = 20 (74.1)<br>AD = 1 (3.7) |
| Amyloid status n (%) | AB + = 117 (29.8)<br>AB - = 271 (68.9)<br>NA = 5 (1.3) | AB + = 147 (31.6)<br>AB - = 310 (66.6)<br>NA = 8 (1.7) | AB + = 264 (30.7)<br>AB - = 581 (67.7)<br>NA = 13 (1.5) | AB + = 20 (100) | AB + = 24 (88.9)<br>AB - = 2 (7.4)<br>NA = 1 (3.7) |

To understand the relationship between tau SUVR and LI, we conducted a univariate correlation between whole temporal SUVR and the absolute value of the LI of the whole temporal lobe (**Figure 1**). We also examined associations between LI with tau SUVR from each individual temporal ROI (**Supplementary Figure 6**), as well as the direct association between left SUVR and right SUVR within each individual temporal ROI (**Supplementary Figure 7**).

**Figure 1:**
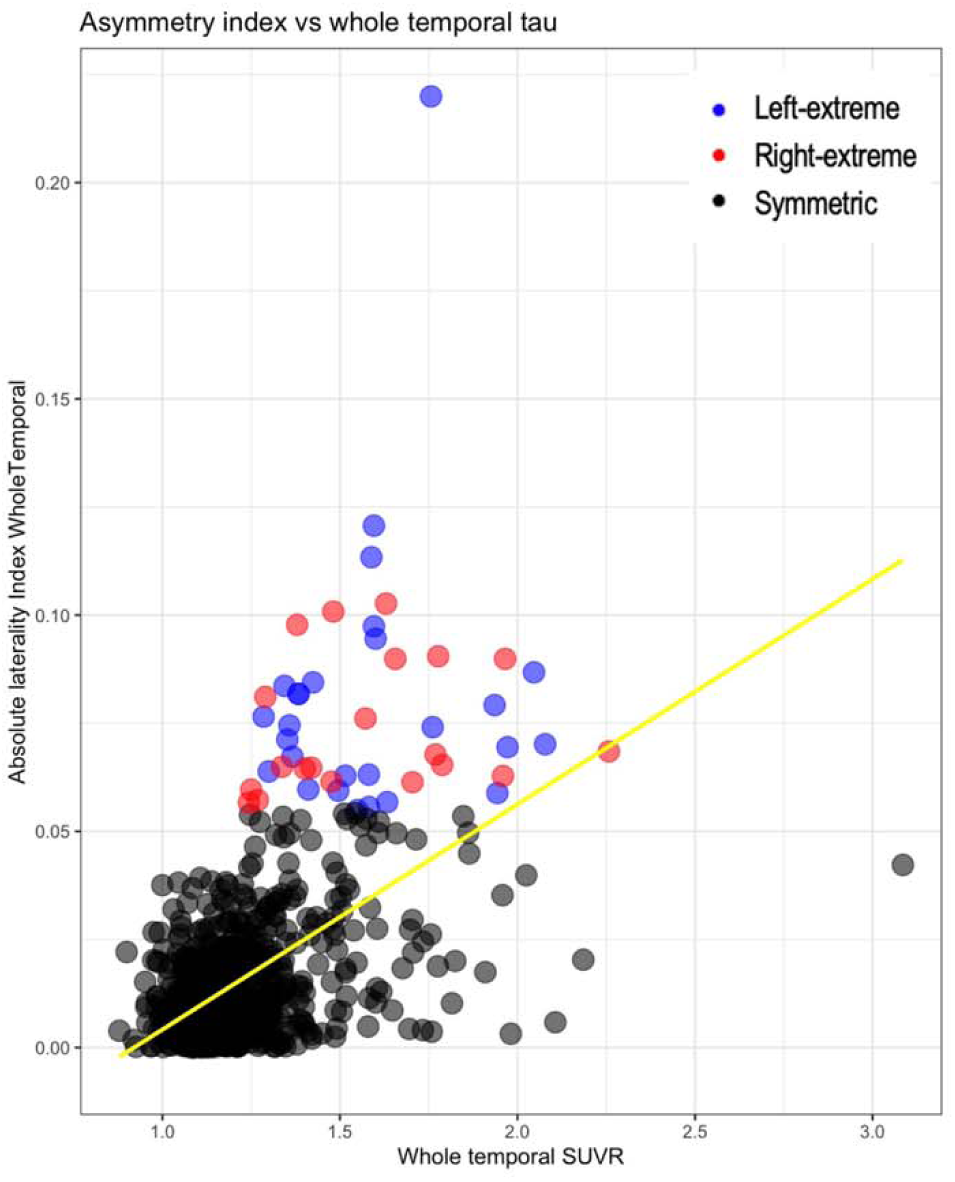
The relationship between whole temporal tau SUVR and absolute laterality index values. Higher whole temporal SUVR is associated with higher temporal tau asymmetry. Asymmetry-extreme cases, colored in blue (left extreme) and red (right extreme), show elevated SUVR values in a range overlapping with the symmetric cases. Yellow line represents regression line for the entire sample.

To determine whether lateralized temporal tau burden was associated with cognitive and behavioral performance, we executed a series of linear regression models within the asymmetry-extreme groups. First, controlling for age, sex, and education, we examined the effect of extreme laterality group (left versus right) on cognitive composite scores and behavior (NPI total score). Next, we examined whether the association between cognition/behavior and elevated tau burden differed by laterality group by including an interaction term between whole temporal SUVR and laterality group into these models.

Leveraging the entire cohort, we examined associations between continuous levels of temporal tau asymmetry with cognitive and behavioral performance. In this analysis, the contribution of each lateralized ROI was examined while controlling for regional tau SUVR in the contralateral ROI. In these models, cognitive composite scores and behavior were the outcome variable, with left and right regional SUVRs are predictors (controlling for age, sex, and education).

## Results

Among the entire sample of 858 subjects, 465 (54%) were on the left lateralized spectrum (LI>0) whereas 393 (46%) were on the right lateralized spectrum (LI<0) (**Figure 1** **and Table 1**). There were no differences in age, education, sex, handedness, race/ethnicity, APOE4, CDR, functional diagnosis, or amyloid status between the two spectrum groups. In the extreme asymmetry groups, there were twenty-seven extreme left and twenty extreme right participants. There were no differences in age, education, sex, CDR-SOB, amyloid status or APOE4 between the left and right extreme groups. The prevalence of left handedness in the right-predominant group was comparable to the 10% prevalence reported in the general population,^33^ whereas the prevalence was relatively higher in the left-predominant group (18% of the left-predominant group were left-handed). Higher whole temporal SUVR was associated with higher temporal tau asymmetry **(Figure 1, Supplementary Figures 6 and 7)**, confirming that asymmetric cases had elevated SUVR values. Further, the range of elevated whole brain and whole temporal values overlapped with the symmetric cases. There were no autosomal dominant mutations careers in the entire cohort.

### Extreme tau asymmetry group confers distinct cognitive and behavioral patterns

The extreme asymmetry groups did not exhibit differences in terms of behavior, memory, executive function, or language after adjusting for age, sex, and education (**Box plots in** **Figure 2A** **and detailed in Table 2 model 1**). Upon the incorporation of whole temporal tau into the model, higher tau was associated with poorer memory and executive performance; however, no associations were observed with behavior or language (**Table 2 model 2**).

**Figure 2:**
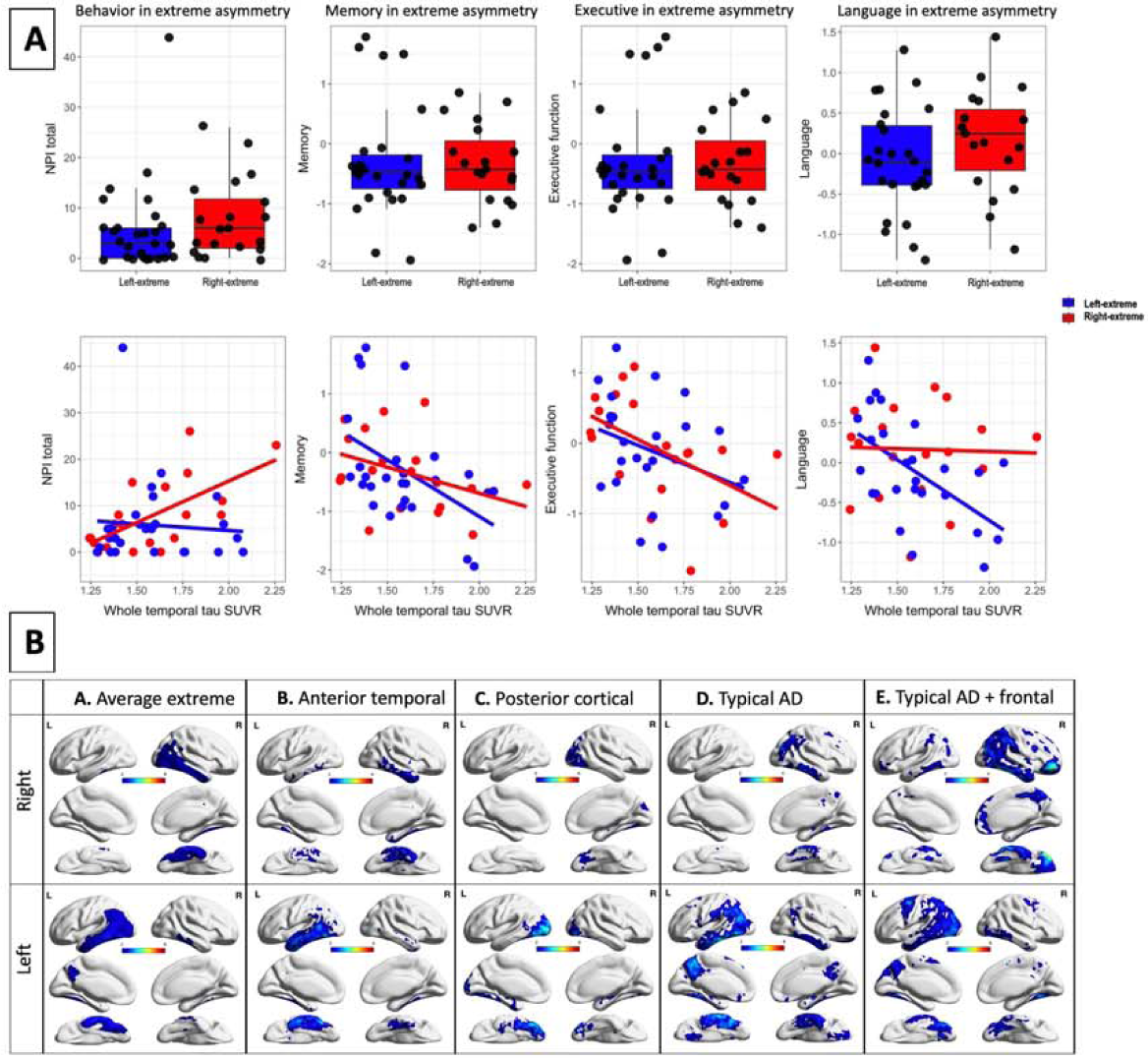
Asymmetry-extreme group. **A-Boxplots:** The asymmetry-extreme groups did not exhibit statistically significant differences on terms of behavior, memory, executive function, or language, although a trend is present for higher NPI in the right-extreme and lower language in the left-extreme. **A-Scatterplots:** There is a significant interaction between whole temporal tau by the right vs left extreme groups; higher right, but not left, tau is associated with worse behavior whereas higher left tau, not right, was associated with worse language symptoms. These distinctions were not present when predicting memory or executive functioning as higher right and left temporal tau were associated with worse memory and executive function. **B.** The first raw represents conjunction maps of the right-extreme group and left-extreme group. Figures were created using tau SUVR threshold of 0.25 (i.e., masks for tau SUVR values over 0.25 were created, binarized, and summed to create conjunction maps). The remaining rows are individual examples from each of the four patterns found upon visual inspection of the asymmetry-extreme groups. The four patterns of asymmetrical tau distribution along the anterior-posterior dimension are shown in B-B. anterior temporal (total of 6 right and 8 left), B-C. posterior (occipital) group (total of 3 right and 1 left) B-D. typical pattern (total of 6 right and 14 left), and B-E. typical pattern with frontal involvement (total of 3 right and 4 left).

**Table.2:** Series of linear regression models in the asymmetry-extreme groups (models 1-3) and in the asymmetry-spectrum (model 4). **Models 1:** The asymmetry-extreme groups did not exhibit differences in terms of behavior, memory, executive function, or language after adjusting for age, sex, and education. **Model 2:** Upon the incorporation of whole temporal tau into the model, higher tau was associated with poorer memory and executive performance; however, no associations were observed with behavior or language. **Model 3:** Upon including the interaction of whole temporal tau by the right vs left extreme groups, higher right, but not left, tau was associated with worse behavior and higher left tau, not right, was associated with worse language symptoms. Model 4 shows the contribution of one ROI after correcting for demographics and contralateral tau. **Model 4:** In the entire cohort, when considering the contralateral tau effect, encompassing both right and left ROIs in the same models and correcting for age, sex, education, chiefly right ROIs displayed significant associations with behavior, while left ROIs were associated with language. Interestingly, more lateral temporal regions were significant (inferior temporal, middle temporal, superior temporal, fusiform gyrus) whereas not all the mesial temporal regions were significant (i.e., medial temporal lobe was trending for behavior on the right and not significant for language on the left). Although the whole temporal, medial temporal lobe, amygdala, and anterior temporal ROIs showed association between higher tau on the left, but not the right, and worse memory and executive function, many temporal ROIs (fusiform gyrus, inferior temporal, middle temporal, and entorhinal cortex displayed correlations with worse both memory and executive functions. NPI: neuropsychiatric inventory. Bold indicates p value less than 0.05. Italics indicate p value less than 0.

| Model 1: Cognition/Behavior ~ Age + Sex + Education + extreme asymmetry group |  |  |  |  |  |  |  |  |
| --- | --- | --- | --- | --- | --- | --- | --- | --- |
|  | NPI |  | Memory |  | Executive |  | Language |  |
| Age | 0.77 (1.2), 0.55 |  | -0.002 (0.1),0.97 |  | -0.04 (0.1), 0.61 |  | -0.02 (0.1), 0.76 |  |
| Education | -0.11 (1.3), 0.93 |  | -0.11 (0.1), 0.20 |  | -0.004 (0.1), 0.96 |  | -0.06 (0.1), 0.49 |  |
| Sex | 0.09 (2.8), 0.97 |  | 0.04 (0.1), 0.82 |  | 0.02 (0.02), 0.92 |  | 0.02 (0.2), 0.89 |  |
| Group (right versus left) | 1.13 (2.6), 0.66 |  | 0.09 (0.1), 0.59 |  | 0.16 (0.2), 0.3 |  | 0.35 (0.17), 0.051 |  |
| Model 2: Cognition/Behavior ~ Age + Sex + Education + extreme asymmetry group + whole temporal tau |  |  |  |  |  |  |  |  |
|  | NPI |  | Memory |  | Executive |  | Language |  |
| Age | 1.39 (1.4), 0.31 |  | -0.07 (0.1), 0.38 |  | -0.12 (0.1), 0.19 |  | -0.07 (0.09), 0.41 |  |
| Education | -0.05 (1.3), 0.96 |  | -0.11 (0.1), 0.16 |  | -0.009 (0.09), 0.91 |  | -0.06 (0.08), 0.46 |  |
| Sex | -0.18 (2.85), 0.94 |  | 0.07 (0.2), 0.68 |  | 0.05 (0.19), 0.76 |  | 0.05 (0.2), 0.79 |  |
| Group (right versus left) | 0.88 (2.6), 0.73 |  | 0.13 (0.16), 0.43 |  | 0.19 (0.17), 0.27 |  | 0.37 (0.17), 0.03 |  |
| Whole Temporal Tau | 1.78 (1.39), 0.21 |  | -0.21 (0.08), 0.02 |  | -0.21 (0.09), 0.02 |  | -0.13 (0.09), 0.14 |  |
| Model 3: Cognition/Behavior ~ Age + Sex + Education + extreme asymmetry group (left vs right) + whole temporal tau + extreme asymmetry group (left vs right) * whole temporal tau |  |  |  |  |  |  |  |  |
|  | NPI |  | Memory |  | Executive |  | Language |  |
| Age | 1.15 (1.3), 0.39 |  | -0.13 (0.1), 0.27 |  | -0.15 (0.1), 0.17 |  | -0.13 (0.1), 0.18 |  |
| Education | -0.07 (1.2), 0.95 |  | -0.001 (0.1), 0.99 |  | -0.05 (0.1), 0.60 |  | -0.01 (0.1), 0.89 |  |
| Sex | -2.06 (2.5), 0.42 |  | 0.49 (0.2), 0.03 |  | 0.37 (0.2), 0.08 |  | 0.27 (0.2), 0.14 |  |
| Group | 1.14 (2.5), 0.65 |  | 0.08 (0.22), 0.59 |  | 0.16 (0.2), 0.44 |  | 0.35 (0.2), 0.06 |  |
| Whole Temporal Tau | -0.28 (1.8), 0.87 |  | -0.51 (0.16), 0.002 |  | -0.31 (0.1), 0.03 |  | -0.43 (0.1), 0.002 |  |
| Group x Whole Temporal Tau | 5.13 (2.4), 0.04 |  | 0.25 (0.22), 0.26 |  | -0.04 (0.2), 0.83 |  | 0.37 (0.1), 0.04 |  |
| Model 4: Cognition/Behavior~ Age+Sex+Education + Right ROI+ Left ROI (R and L tau ROIs in same model) |  |  |  |  |  |  |  |  |
|  | NPI |  | Memory |  | Executive |  | Language |  |
|  | R | L | R | L | R | L | R | L |
| Whole temporal | 7.19 (2.9), 0.01 | 1.41 (2.86), 0.62 | -0.43 (0.27), 0.112 | -1.41 (0.25), <.0001 | -0.42 (0.26), 0.11 | -0.9 (0.25), .0001 | 0.31 (0.25), 0.20 | -1.40 (0.24), <.0001 |
| Fusiform | 5.12 (1.89), 0.007 | 0.75 (1.75), 0.66 | -0.37 (0.17), 0.03 | -0.91 (0.16), <.0001 | -0.27 (0.16), 0.10 | -0.79 (0.15), <.0001 | 0.10 (0.16), 0.53 | -0.92 (0.15), <.0001 |
| Inferior temporal | 4.52 (1.62), 0.005 | 0.82 (1.52), 0.58 | -0.29 (0.14), 0.04 | -0.86 (0.13), <.0001 | -0.39 (0.14), 0.005 | -0.59 (0.13), <.0001 | -0.001 (0.13), 0.99 | -0.76 (0.12), <.0001 |
| Middle temporal | 4.07 (1.62), 0.01 | 1.62 (1.53), 0.29 | -0.54 (0.14), 0.0002 | -0.7 (0.14), <.0001 | -0.47 (0.14), 0.0008 | -0.59 (0.13), 0.0008 | -0.08 (0.13), 0.51 | -0.77 (0.12), <.0001 |
| Superior temporal | 6.11 (2.69), 0.02 | 1.22 (2.40), 0.61 | -0.77 (0.25), 0.002 | -0.97 (0.23), <.0001 | -0.59 (0.24), 0.01 | -0.84 (0.21), 0.0001 | 0.07 (0.22), 0.74 | -1.20 (0.20), <.0001 |
| Anterior temporal | 2.11 (2.57), 0.41 | 5.95 (2.33), 0.01 | -0.36 (0.24), 0.12 | -1.28 (0.21), <.0001 | -0.03 (0.23), 0.13 | -0.79 (0.21), <.0002 | -0.001 (0.21), 0.99 | -1.01 (0.19), <.0001 |
| Amygdala | 6.59 (2.95), 0.02 | 1.38 (2.89), 0.63 | -0.09 (0.27), 0.73 | -1.45 (0.26), <.0001 | -0.27 (0.27), 0.32 | -0.67 (0.27), 0.01 | 0.19 (0.26), 0.46 | -0.89 (0.25), 0.0005 |
| Entorhinal cortex | 5.1 (2.4), 0.03 | 2.65 (2.41), 0.27 | -0.49 (0.22), 0.02 | -1.14 (0.22), <.0001 | -0.63 (0.22), 0.004 | -0.47, (0.22), 0.03 | 0.04 (0.2), 0.84 | -0.9 (0.21), <.0001 |
| Medial temporal lobe | 6.99 (3.62), 0.053 | 2.13 (3.58), 0.55 | -1.31 (0.33), 0.34 | -1.54 (0.32), <.0001 | -0.47 (0.33), 0.15 | -0.75 (0.33), 0.02 | -1.16 (0.31), .0002 | 0.23 (0.31), 0.45 |

An interaction term between laterality group and whole temporal tau SUVR showed distinct patterns with behavior and cognition within each group. Specifically, the right temporal extreme asymmetry group displayed increased behavioral symptoms as tau burden increased, while language symptoms remained relatively stable with increasing tau levels. On the other hand, the left temporal extreme asymmetry group exhibited lower language scores as tau burden increased, while behavioral symptoms remained relatively steady with rising tau levels (**Scatter plots in** **Figure 2A** **and detailed in Table 2 model 3**). These distinctions were not present when predicting memory or executive functioning.

### NPI subcomponents

Given that right-extreme tau groups showed worse performance on the NPI total score, we further investigated which NPI subcomponents were driving this effect. Anxiety and hallucinations were associated with higher tau and irritability/lability trended towards significance. There were no differences in aberrant motor behavior, disinhibition, apathy/indifference, elation/euphoria, agitation/aggression, delusions, sleep, or appetite and eating disorders (**Supplementary Table 1 and Supplementary Figure 5**).

### Patterns of tau asymmetry across an anterior-posterior axis

Average maps of the right and left asymmetry extreme groups are shown in **Figure 2B-A**. However, visual inspection of individuals within each asymmetry-extreme group revealed at least four patterns of asymmetrical tau distribution along the anterior-posterior dimension. We noted individuals with patterns of tau deposition in a typical AD pattern (**Figure 2B-D**), a typical AD pattern that extended into frontal areas (**Figure 2B-E**), anterior temporal (**Figure 2B-B**), and posterior/occipital (**Figure 2B-C**). Among the right-predominant cases, six (30%) predominantly involved the anterior temporal lobe reminiscent of sbvFTD. Three cases (15%) displayed a right occipital pattern reminiscent of posterior cortical atrophy. Additionally, six cases (30%) exhibited right posterior lateral temporoparietal region involvement, and three cases (15%) showed right posterior lateral temporoparietal region involvement along with frontal region involvement. Similarly, within the left-predominant group, eight subjects (30%) primarily displayed left temporal predominance, mainly encompassing anterior temporal regions (similar to svPPA). One case (4%) resembled a left occipital pattern reminiscent of posterior cortical atrophy. Furthermore, fourteen cases (52%) featured left posterior lateral temporoparietal region involvement, and four cases (15%) presented left posterior lateral temporoparietal region involvement along with frontal region involvement.

### Asymmetry spectrum of tau confers distinct cognitive and behavioral patterns

In the entire cohort, when controlling for contralateral tau, right regional temporal tau levels displayed significant associations with behavior, while left regional temporal tau levels were associated with language (**Table 2**, **Figure 3**, **supplementary Figures 1-4**). Interestingly, these effects were more consistent in lateral temporal regions (inferior temporal, middle temporal, superior temporal, fusiform gyrus) as opposed to medial temporal regions. The relationship between tau laterality was less clear for memory and executive function. Although the whole temporal, medial temporal lobe, amygdala, and anterior temporal ROIs showed associations between higher left asymmetric tau with worse memory and executive function, many other additional ROIs on both the left and right (fusiform gyrus, inferior temporal, middle temporal, and entorhinal cortex) displayed correlations with worse memory and executive function.

**Figure 3.**
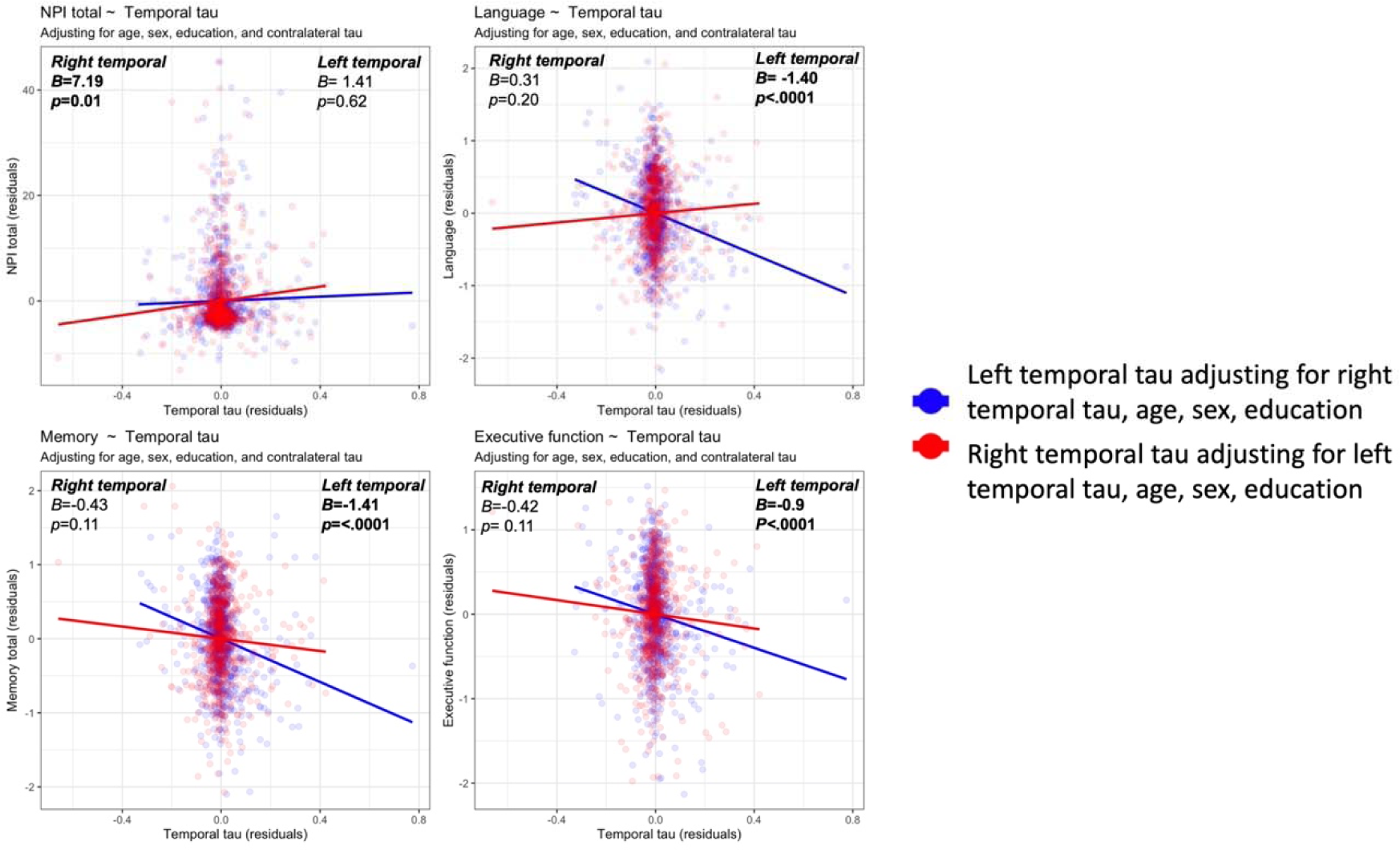
Association of right and left temporal tau with each of behavior as measures by the neuropsychiatric inventory (NPI) and cognition as measured by language, memory, and executive function composites. Figure shows residualized contributions of the left and right temporal ROI after correcting for the contralateral temporal ROI as well as age, sex, and education. Upper left: Higher right-temporal but not left-temporal associated with worse behavioral symptoms. Upper right: Higher left-temporal but not right-temporal associated with worse language performance. Lower left: Higher left-temporal but not right-temporal associated with worse memory performance. Upper right: Higher left-temporal but not right-temporal associated with worse executive function performance. Similar plots are shown for each temporal ROI in Supplementary Materials Figure 1-4.

## Discussion

This study characterized the presence of asymmetric patterns of tau accumulation in a mildly impaired AD cohort with a primarily amnestic presentation. Importantly, the degree of asymmetry within the temporal lobe corresponded to distinct behavioral and language deficits. Predominant right temporal tau accumulation was associated preferentially with behavioral symptoms, whereas predominant left temporal tau accumulation was linked preferentially to language symptoms. Furthermore, patterns of asymmetry were not consistent across individuals, suggesting multiple subtypes exist spanning the anterior-posterior axis. Interestingly, there were less clearly discernible differences between right and left predominant temporal tau groups in memory and executive function. These observations hold implications for clinical diagnosis, disease progression tracking, and therapeutic approaches. Our findings align with the established typical amnestic AD, limbic predominant, and hippocampal sparing schema^12,13^ and extend this classification by noting additional heterogeneity regarding laterality, as well as additional subtypes such as an anterior temporal variant that aligns anatomically with frontotemporal dementia (FTD) subtypes. Altogether, this study highlights the need to integrate and develop additional socioemotional and neuropsychological measures that may be most impacted by tau laterality in the temporal lobes.

The early involvement of the temporal lobes in AD is well-documented. Aside from their role in episodic memory and spatial navigation, the temporal lobes function as central hubs for both verbal and non-verbal semantics. Verbal semantics encompass multi-modal knowledge of words and objects and are preferentially left temporal lateralized. Whereas non-verbal semantics involve multi-modal knowledge for socio-emotionally relevant concepts.^21,34,35^ Extensive FTD, linguistic, and socioemotional literature show that semantic concepts are represented on a graded spectrum where both right and left temporal lobes seem to be required for semantic knowledge with the right being more specialized for non-verbal socioemotional concepts and the left for verbal semantics.^23,36,37^ Over the past couple of decades, research has elucidated the networks responsible for emotion processing. Processing the emotions of others involves several steps including understanding the meaning of an external expression, internally experiencing the expressed emotion, attributing the behavior to the other rather than the self, and inhibiting one’s own perspective.^38,39^ These processes localize to different but interconnected neuroanatomical circuits communicating between the insula, temporo-parietal junction, and the semantic appraisal network connecting the anterior temporal lobe and the orbitofrontal cortex. We presume that all these regions can also be afflicted by AD pathology. Previous studies have indicated that AD patients perform poorly on emotion perception tasks.^40^ In the current study, the left-predominant group exhibited more pronounced language deficits whereas the right-predominant group showed associations with behavioral scores. This pattern of verbal/left and non-verbal right lateralization aligns with the literature in FTD, which indicates a spectrum of semantic loss on a spectrum spanning non-verbal socioemotional concepts in the right and verbal semantic on the left temporal lobes. Collectively, we propose that the interaction between tau and the language network in the left, and with the non-verbal socioemotional network in the right, could potentially be caused by the loss of verbal and non-verbal semantics, respectively.^21^

Behavioral symptoms in AD in relation to frontal cortex are relevant to a previously described variant labeled executive, or behavioral variant AD.^41–47^ Comparisons between typical amnestic AD and behavioral variant AD point to divergent patterns of cortical atrophy,^41,42^ glucose metabolism,^43^ and tau accumulation.^48^ However, these studies have limitations such as not including measures of behavior or considering both hemispheres simultaneously. Furthermore, pathological studies often fail to determine whether the right or left hemispheres are simultaneously or asymmetrically involved, mainly because they typically focus on examining one hemisphere. Additionally, in-vivo tau PET imaging studies have widely utilized a meta-temporal ROI to estimate tau burden,^44–47^ which combines regions in the temporal lobes and collapsed across hemisphere. In our study, we demonstrate that even in a typical amnestic cohort of AD, there is asymmetric tau accumulation associated with distinct phenotypic patterns along the behavioral/language continuum. Consistent with the right temporal role in emotions in AD, research investigating cortical volumes showed that emotional contagion is linked to atrophy in the right lateral temporal regions.^22^ Moreover, reduced emotion prosody recognition has been associated with cortical atrophy in the right temporal pole and superior temporal sulcus in AD.^49^ Furthermore, right medial temporal lobe involvement has been linked to paranoid delusions in mild AD.^50^ In our study, the differences observed in the NPI were primarily influenced by increased anxiety, irritability/liability, and hallucinations. Intriguingly, a functional MRI investigation revealed a connection between heightened salience network connectivity in the anterior cingulate cortex and right insula areas, and specific NPI subcomponents including agitation, irritability, aberrant motor behavior, euphoria, and disinhibition.^51^

When examined individually, the right- and left-extreme participants exhibited variable degrees of frontal, parietal, and occipital involvement. When defining these extreme asymmetry groups, we did not restrict selection to cases that only had asymmetry in the right or left temporal lobe and it is possible that a given subject’s maximum tau accumulation was outside the temporal lobes. Further, the observed patterns of variability along the anterior-posterior axis adds to the complexity of phenotypic and pathological heterogeneity. Interestingly, the left-predominant group involved more of the contralateral hemisphere compared to the right-predominant group consistent with research showing cortical volume loss occurring earlier and progressing faster in the left hemisphere.^52,53^ However, this may be a confound related to study design. Specifically, participants are recruited based on language centered tests and must meet ADNI mild cognitive impairment or dementia criteria for enrollment whereas advanced right predominant cases may be excluded due to more pronounced behavioral symptoms that are presumed to be due to non-AD etiologies. Additionally, the neuropsychological tools that evaluate memory and executive function are language-dependent biasing sample selection even further to left lateralized disease and highlighting the need for non-verbal neuropsychological batteries to understand the full phenotypic heterogeneity associated with AD pathology.

Mechanisms underlying asymmetry and heterogeneity in the spatial distribution of tau in AD remain unknown. Asymmetric patterns in AD are not specific to PET and have been found in cortical volumes,^49^ white matter topology,^54^ and structural connectivity networks.^50^ Furthermore, early pathological investigations reported asymmetric hemispheric, hippocampal plaques and tangles, and lateralized differences within Braak and Braak stages in at least 15% of cases.^56–58^ Gaining a better understanding of asymmetry in AD could offer insights into the underlying causes of selective vulnerability.

This study has limitations. It was performed in the ADNI cohort which primarily consists of amnestic AD. Consequently, the study’s findings likely underestimate the prevalence or degree of right-predominant patterns in the broader population. It is important to note that large cohorts with a broad range of clinical symptoms that have undergone tau PET are currently lacking. A notable limitation is the absence of in depth objective measures for assessing behavior, emotion, and language in ADNI. Our behavioral analyses rely on the NPI questionnaire, which likely shows limited sensitivity to detecting right-sided deficits in behavior and emotion.

In conclusion, this study highlights the importance of asymmetry in patterns of tau burden in AD patients. It is imperative to develop better behavioral, socioemotional, and language tests to understand this phenotypic heterogeneity. Therapeutic and diagnostic approaches in precision medicine would benefit from characterizing the asymmetry burden in the spatial distribution of tau in patients with AD, as this asymmetry has implications for clinical impairment, caregiver burden, and disease trajectories.

## Data availability statement

Data for this study is publicly available on the ADNI LONI website https://adni.loni.usc.edu/

## References

1. Alzheimer A. Uber eigenartige Erkrankung der Hirnrinde. All Z Psychiatr. 1907;64:146–148.

2. Arriagada P V, Growdon JH, Hedley-Whyte ET, Hyman BT. Neurofibrillary tangles but not senile plaques parallel duration and severity of Alzheimer’s disease. Neurology. 1992;42(3 Pt 1):631–9. http://www.ncbi.nlm.nih.gov/htbin-post/Entrez/query?db=m&form=6&dopt=r&uid=1549228

3. Bejanin A, Schonhaut DR, La Joie R, et al. Tau pathology and neurodegeneration contribute to cognitive impairment in Alzheimer’s disease. Brain. 2017;140(12):3286–3300. doi:10.1093/brain/awx243

4. Russ TC, Batty GD, Starr JM. Cognitive and behavioural predictors of survival in Alzheimer disease: Results from a sample of treated patients in a tertiary-referral memory clinic. Int J Geriatr Psychiatry. 2012;27(8):844–853. doi:10.1002/gps.2795

5. Terum TM, Andersen JR, Rongve A, Aarsland D, Svendsboe EJ, Testad I. The relationship of specific items on the Neuropsychiatric Inventory to caregiver burden in dementia: a systematic review. Int J Geriatr Psychiatry. 2017;32(7):703–717. doi:10.1002/gps.4704

6. Bucks RS, Byrne L, Haworth J, et al. The cost of behavioral and psychological symptoms of dementia (BPSD) in community dwelling Alzheimer’s disease patients. Int J Geriatr Psychiatry. 2002;17(5):403–408. doi:10.1002/gps.490

7. Apostolova LG, Cummings JL. Neuropsychiatric manifestations in mild cognitive impairment: a systematic review of the literature. Dement Geriatr Cogn Disord. 2008;25(2):115–126. http://www.ncbi.nlm.nih.gov/entrez/query.fcgi?cmd=Retrieve&db=PubMed&dopt=Citation&list_uids=18087152

8. Spalletta G, Musicco M, Padovani A, et al. Neuropsychiatric symptoms and syndromes in a large cohort of newly diagnosed, untreated patients with Alzheimer disease. Am J Geriatr Psychiatry. 2010;18(11):1026–1035. doi:10.1097/JGP.0b013e3181d6b68d

9. Albert MS, Dekosky ST, Dickson D, et al. The diagnosis of mild cognitive impairment due to Alzheimer’s disease: Recommendations from the National Institute on Aging and Alzheimer’s Association workgroup. Alzheimers Dement. Published online 2011. http://www.ncbi.nlm.nih.gov/entrez/query.fcgi?cmd=Retrieve&db=PubMed&dopt=Citation&list_uids=21514249

10. Braak H, Thal DR, Ghebremedhin E, Del Tredici K. Stages of the pathologic process in Alzheimer disease: age categories from 1 to 100 years. J Neuropathol Exp Neurol. 2011;70(11):960–969. doi:10.1097/NEN.0b013e318232a379

11. Grinberg LT, Rüb U, Ferretti REL, et al. The dorsal raphe nucleus shows phospho-tau neurofibrillary changes before the transentorhinal region in Alzheimer’s disease. A precocious onset? Neuropathol Appl Neurobiol. 2009;35(4):406–416. doi:10.1111/j.1365-2990.2009.00997.x

12. Murray ME, Graff-Radford NR, Ross OA, Petersen RC, Duara R, Dickson DW. with distinct clinical characteristicsD: A retrospective study. Lancet Neurol. 2011;10(9):785–796. doi:10.1016/S1474-4422(11)70156-9.Neuropathologically

13. Whitwell JL, Dickson DW, Murray ME, et al. Neuroimaging correlates of pathologically defined subtypes of Alzheimer’s disease: A case-control study. Lancet Neurol. 2012;11(10):868–877. doi:10.1016/S1474-4422(12)70200-4

14. Vogel JW, Young AL, Oxtoby NP, et al. Four distinct trajectories of tau deposition identified in Alzheimer’s disease. Nat Med. 2021;27(5):871–881. doi:10.1038/s41591-021-01309-6

15. Dong A, Toledo JB, Honnorat N, et al. Heterogeneity of neuroanatomical patterns in prodromal Alzheimer’s disease: links to cognition, progression and biomarkers. Brain. 2017;140(3):735–747. doi:10.1093/brain/aww319

16. Ossenkoppele R, Lyoo CH, Sudre CH, et al. Distinct tau PET patterns in atrophy-defined subtypes of Alzheimer’s disease. Alzheimers Dement. 2020;16(2):335–344. doi:10.1016/j.jalz.2019.08.201

17. Young CB, Winer JR, Younes K, et al. Divergent Cortical Tau Positron Emission Tomography Patterns Among Patients With Preclinical Alzheimer Disease. JAMA Neurol. 2022;79(6):592–603. doi:10.1001/jamaneurol.2022.0676

18. Noh Y, Lee JM, Kim GH, et al. Anatomical heterogeneity of Alzheimer disease Based on cortical thickness on MRIs. Published online 2014.

19. Younes K, Miller BL. Frontotemporal Dementia: Neuropathology, Genetics, Neuroimaging, and Treatments. Psychiatr Clin North Am. Published online 2020. doi:10.1016/j.psc.2020.02.006

20. Gorno-Tempini ML, Hillis AE, Weintraub S, et al. Classification of primary progressive aphasia and its variants. Neurology. 2011;76(11):1006–1014. doi:10.1212/WNL.0b013e31821103e6

21. Younes K, Borghesani V, Montembeault M, et al. Right temporal degeneration and socioemotional semantics: semantic behavioural variant frontotemporal dementia. Brain. 2022;145(11):4080–4096. doi:10.1093/brain/awac217

22. Sturm VE, Yokoyama JS, Seeley WW, Kramer JH, Miller BL, Rankin KP. Heightened emotional contagion in mild cognitive impairment and Alzheimer’s disease is associated with temporal lobe degeneration. Proc Natl Acad Sci U S A. 2013;110(24):9944–9949. doi:10.1073/pnas.1301119110

23. Hoffman P, Jones RW, Ralph MAL. The degraded concept representation system in semantic dementia: Damage to pan-modal hub, then visual spoke. Brain. 2012;135(12):3770–3780. doi:10.1093/brain/aws282

24. Younes K, Miller BL. Frontotemporal Dementia: Neuropathology, Genetics, Neuroimaging, and Treatments. Psychiatr Clin North Am. 2020;43(2):331–344. doi:10.1016/j.psc.2020.02.006

25. Younes K, Miller BL. Neuropsychiatric Aspects of Frontotemporal Dementia. Psychiatr Clin North Am. Published online 2020. doi:10.1016/j.psc.2020.02.005

26. Borghesani V, Battistella G, Mandelli ML, et al. Regional and hemispheric susceptibility of the temporal lobe to FTLD-TDP-43-C pathology. bioRxiv. Published online 2020:847582. doi:10.1101/847582

27. Morris JC. The clinical dementia rating (cdr): Current version and scoring rules. Neurology. 1993;43(11):2412–2414. doi:10.1212/wnl.43.11.2412-a

28. Mukherjee S, Choi SE, Lee ML, et al. Cognitive Domain Harmonization and Cocalibration in Studies of Older Adults. Neuropsychology. Published online 2022. doi:10.1037/neu0000835

29. Cummings JL. The Neuropsychiatric Inventory: assessing psychopathology in dementia patients. Neurology. 1997;48(5 Suppl 6):S10–S16. doi:10.1212/wnl.48.5_suppl_6.10s

30. Rabinovici GD, Jagust WJ, Furst AJ, et al. Abeta amyloid and glucose metabolism in three variants of primary progressive aphasia. Ann Neurol. 2008;64(4):388–401. http://www.ncbi.nlm.nih.gov/entrez/query.fcgi?cmd=Retrieve&db=PubMed&dopt=Citation&list_uids=18991338

31. Borghesani V, Battistella G, Mandelli ML, et al. Regional and hemispheric susceptibility of the temporal lobe to FTLD-TDP type C pathology. NeuroImage Clin. 2020;28(November 2019):102369. doi:10.1016/j.nicl.2020.102369

32. Landau SM, Ward TJ, Murphy A, et al. Quantification of amyloid beta and tau PET without a structural MRI. Alzheimer’s Dement. 2022;2022(February):1–12. doi:10.1002/alz.12668

33. McManus C. Half a century of handedness research: Myths, truths; fictions, facts; backwards, but mostly forwards. Brain Neurosci Adv. 2019;3:239821281882051. doi:10.1177/2398212818820513

34. Ralph MAL, Jefferies E, Patterson K, Rogers TT. The neural and computational bases of semantic cognition. Nat Rev Neurosci. 2017;18(1):42–55. doi:10.1038/nrn.2016.150

35. Binney RJ, Ramsey R. Controlled Social Cognition: the role of conceptual knowledge and cognitive control in a neurobiological model of the social brain. PsyArXiv Prepr. Published online 2019:1–45.

36. Hodges JR, Patterson K, Oxbury S, Funnell E. Semantic dementia. Progressive fluent aphasia with temporal lobe atrophy. Brain. 1992;115(Pt 6):1783–1806.

37. Snowden JS, Harris JM, Thompson JC, et al. Semantic dementia and the left and right temporal lobes. Cortex. 2018;107(September):188–203. doi:10.1016/j.cortex.2017.08.024

38. Rankin Katherine. P., Gorno-Tempini Luisa. M, Stephen. C. A, et al. Structural anatomy of empathy in neurodegenerative disease. Brain. 2006;129(11):2945–2956. doi:10.1093/brain/awl254

39. Decety J, Jackson PL. The functional architecture of human empathy. Behav Cogn Neurosci Rev. 2004;3(2):71–100. doi:10.1177/1534582304267187

40. Elferink MWO, Van Tilborg I, Kessels RPC. Perception of emotions in mild cognitive impairment and Alzheimer’s dementia: Does intensity matter? Transl Neurosci. 2015;6(1):139–149. doi:10.1515/tnsci-2015-0013

41. Ossenkoppele R, Pijnenburg YA, Perry DC, et al. The behavioural/dysexecutive variant of Alzheimer’s disease: clinical, neuroimaging and pathological features. Brain. 2015;138(Pt 9):2732–2749. doi:10.1093/brain/awv191

42. Ossenkoppele R, Singleton EH, Groot C, et al. Research Criteria for the Behavioral Variant of Alzheimer Disease: A Systematic Review and Meta-analysis. JAMA Neurol. 2022;79(1):48–60. doi:10.1001/jamaneurol.2021.4417

43. Sala A, Caprioglio C, Santangelo R, et al. Brain metabolic signatures across the Alzheimer’s disease spectrum. Eur J Nucl Med Mol Imaging. 2020;47(2):256–269. doi:10.1007/s00259-019-04559-2

44. Schöll M, Lockhart SN, Schonhaut DR, et al. PET Imaging of Tau Deposition in the Aging Human Brain. Neuron. 2016;89(5):971–982. doi:10.1016/j.neuron.2016.01.028

45. Edwards L, La Joie R, Iaccarino L, et al. Multimodal neuroimaging of sex differences in cognitively impaired patients on the Alzheimer’s continuum: greater tau-PET retention in females. Neurobiol Aging. 2021;105:86–98. doi:10.1016/j.neurobiolaging.2021.04.003

46. Cho H, Choi JY, Hwang MS, et al. In vivo cortical spreading pattern of tau and amyloid in the Alzheimer disease spectrum. Ann Neurol. 2016;80(2):247–258. doi:10.1002/ana.24711

47. Mormino EC, Toueg TN, Azevedo C, et al. Tau PET imaging with 18F-PI-2620 in aging and neurodegenerative diseases. Eur J Nucl Med Mol Imaging. 2021;48(7):2233–2244. doi:10.1007/s00259-020-04923-7

48. Singleton E, Hansson O, Pijnenburg YAL, et al. Heterogeneous distribution of tau pathology in the behavioural variant of Alzheimer’s disease. J Neurol Neurosurg Psychiatry. 2021;92(8):872–880. doi:10.1136/jnnp-2020-325497

49. Amlerova J, Laczó J, Nedelska Z, et al. Emotional prosody recognition is impaired in Alzheimer’s disease. Alzheimer’s Res Ther. 2022;14(1):1–8. doi:10.1186/s13195-022-00989-7

50. Geroldi C, Akkawi NM, Galluzzi S, et al. Temporal lobe asymmetry in patients with Alzheimer’s disease with delusions. J Neurol Neurosurg Psychiatry. 2000;69(2):187–191. doi:10.1136/jnnp.69.2.187

51. Balthazar MLF, Pereira FRS, Lopes TM, et al. Neuropsychiatric symptoms in Alzheimer’s disease are related to functional connectivity alterations in the salience network. Hum Brain Mapp. 2014;35(4):1237–1246. doi:10.1002/hbm.22248

52. Janke AL, De Zubicaray G, Rose SE, Griffin M, Chalk JB, Galloway GJ. 4D deformation modeling of cortical disease progression in Alzheimer’s dementia. Magn Reson Med. 2001;46(4):661–666. doi:10.1002/mrm.1243

53. Thompson PM, Hayashi KM, de Zubicaray G, et al. Dynamics of gray matter loss in Alzheimer’s disease. J Neurosci. 2003;23(3):994–1005. http://www.ncbi.nlm.nih.gov/entrez/query.fcgi?cmd=Retrieve&db=PubMed&dopt=Citation&list_uids=12574429

54. Daianu M, Jahanshad N, Nir TM, et al. Breakdown of brain connectivity between normal aging and Alzheimer’s disease: A structural k-Core network analysis. Brain Connect. 2013;3(4):407–422. doi:10.1089/brain.2012.0137

55. Roe JM, Vidal-Piñeiro D, Sørensen Ø, et al. Asymmetric thinning of the cerebral cortex across the adult lifespan is accelerated in Alzheimer’s disease. Nat Commun. 2021;12(1):1–11. doi:10.1038/s41467-021-21057-y

56. Stefanits H, Budka H, Kovacs GG. Asymmetry of neurodegenerative disease-related pathologies: A cautionary note. Acta Neuropathol. 2012;123(3):449–452. doi:10.1007/s00401-011-0936-6

57. Hyman BT, Phelps CH, Beach TG, et al. National Institute on Aging-Alzheimer’s Association guidelines for the neuropathologic assessment of Alzheimer’s disease. Alzheimer’s Dement. 2012;8(1):1–13. doi:10.1016/j.jalz.2011.10.007

58. King A, Bodi I, Nolan M, Troakes C, Al-Sarraj S. Assessment of the degree of asymmetry of pathological features in neurodegenerative diseases. What is the significance for brain banks? J Neural Transm. 2015;122(10):1499–1508. doi:10.1007/s00702-015-1410-8

