## Supplementary Materials for "Temporal tau asymmetry spectrum influences divergent behavior and language patterns in Alzheimer’s disease"

**Methods:**

**Alzheimer`s Disease Neurogimaing Initiaitve (ADNI):** The ADNI is a public-private partnership launched in 2003 with the primary goal of testing whether serial neuroimaging and biological markers, and clinical and neuropsychological assessments can be combined to measure the progression of MCI and early AD. For up-to-date information, see [www.adni-info.org](http://www.adni-info.org).

**The clinical dementia rating scale (CDR):** The CDR characterizes six domains of cognitive and functional performance including Memory, Orientation, Judgment & Problem Solving, Community Affairs, Home & Hobbies, and Personal Care. The sum of boxes of the CDR (CDR-SB) is the sum score of the six domains.

**Genetic data in ADNI:** Various types of genetic data are available in ADNI, including APOE, TOMM40 poly-T repeat, GWAS, whole exome sequences, whole genome sequences, and microarray-based RNA gene expression profiles.

**Supplementary figures:**

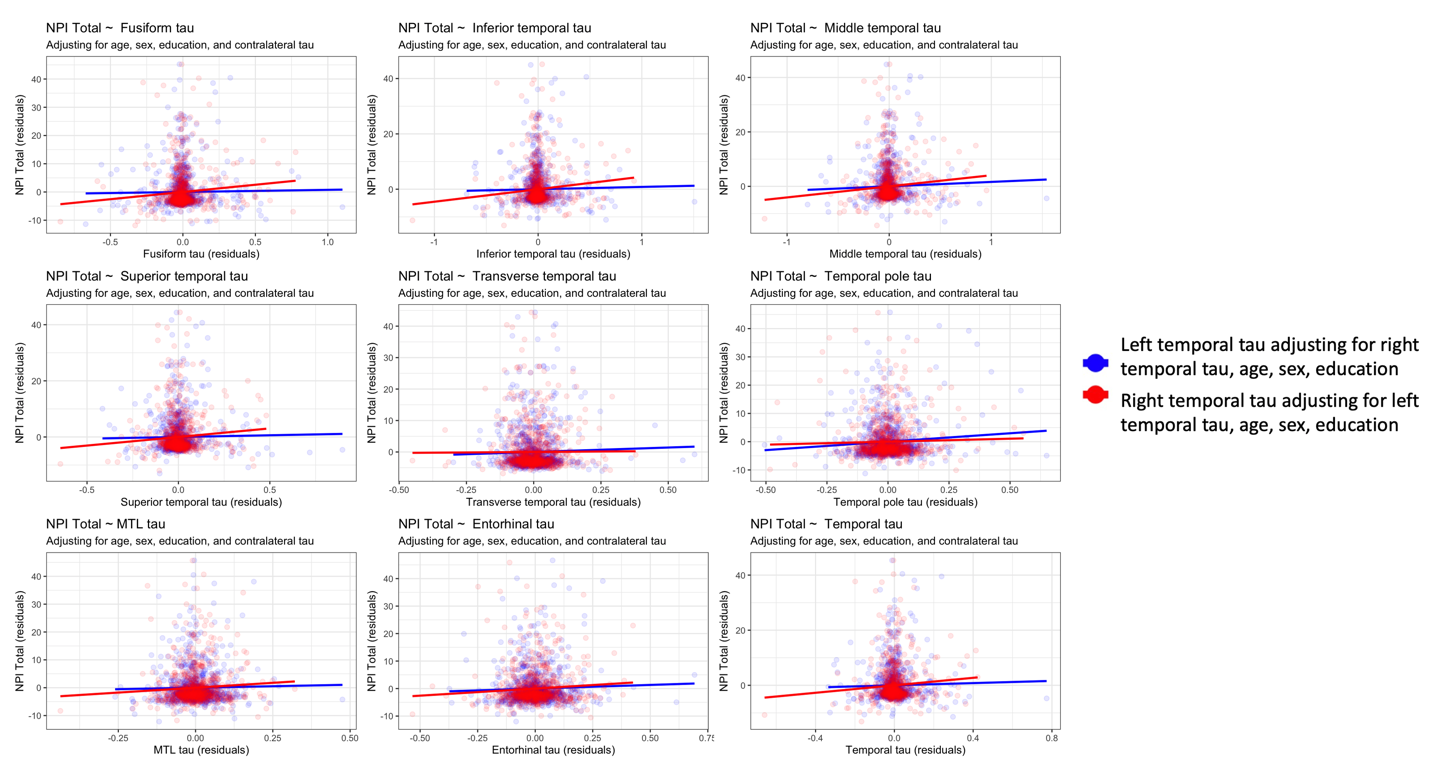

**Supplementary Figure.1.** Association of right and left temporal tau ROIs with behavior as measures by the neuropsychiatric inventory (NPI).

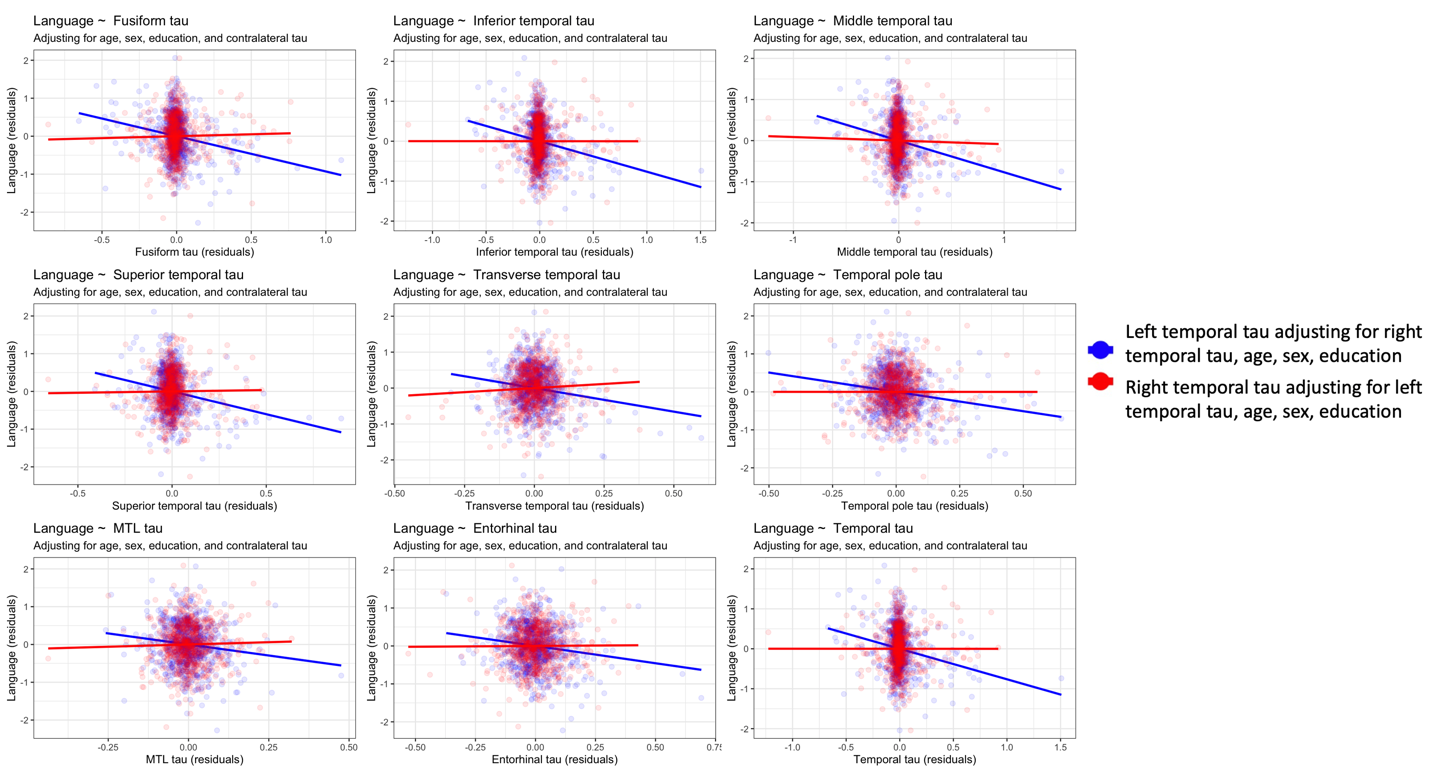

**Supplementary Figure.2.** Association of right and left temporal tau ROIs with language composite.

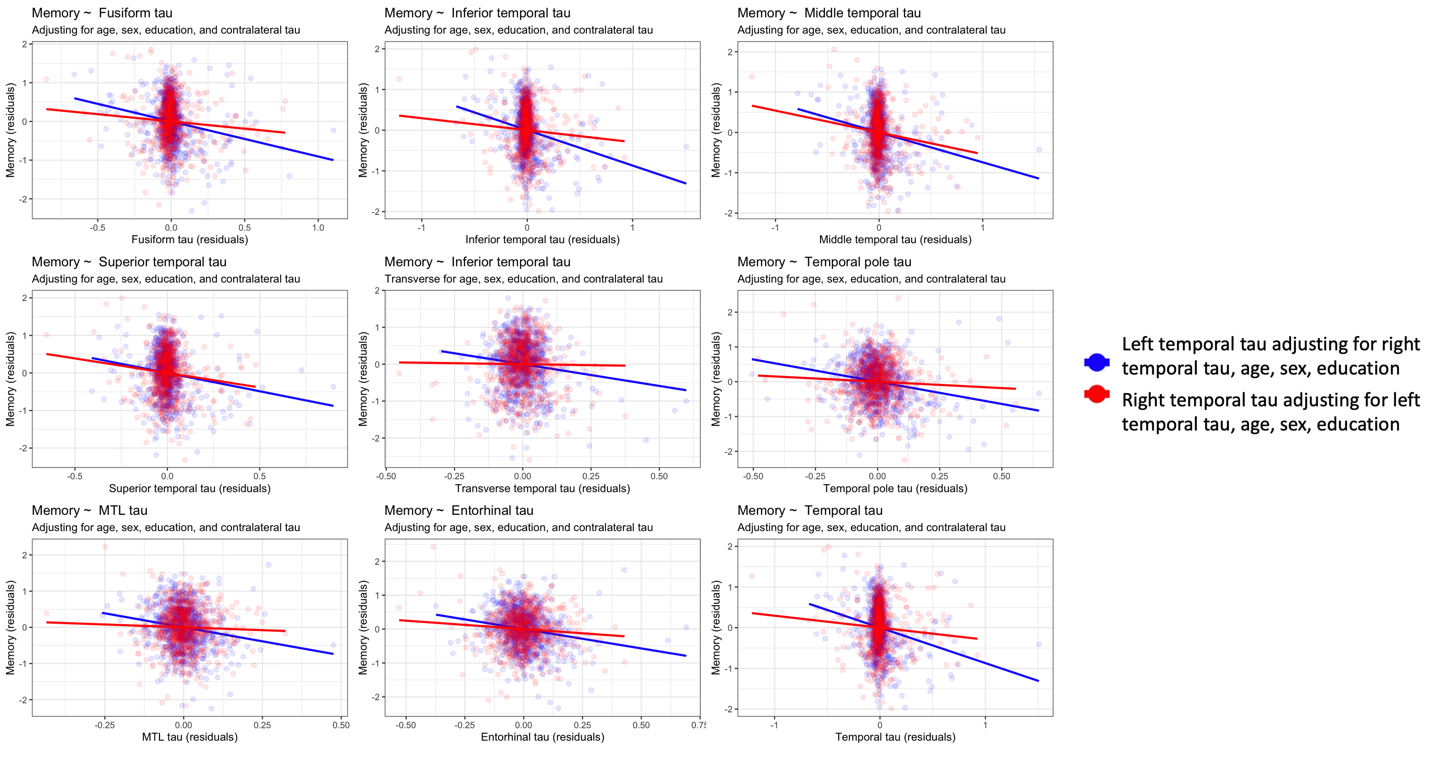

**Supplementary Figure.3.** Association of right and left temporal tau ROIs with memory composite.

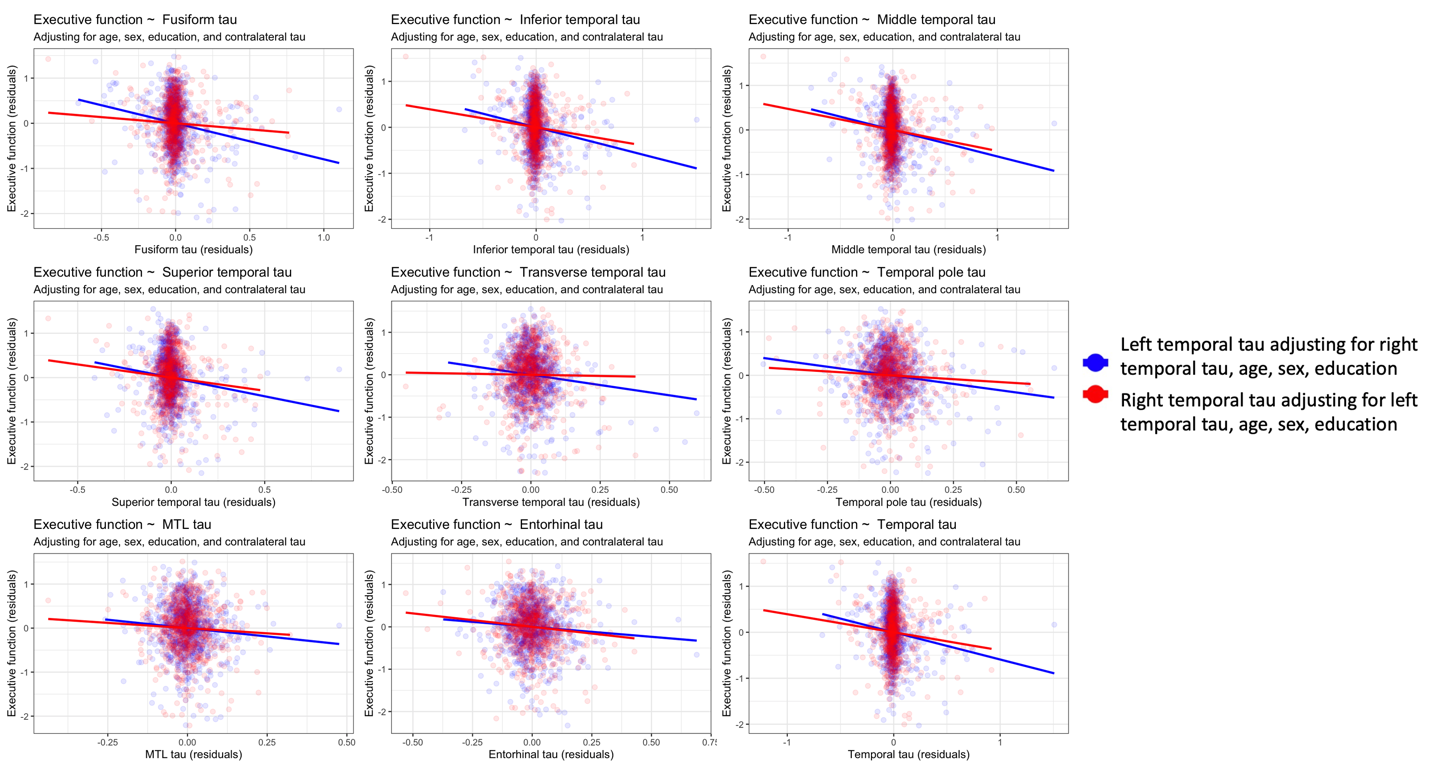

**Supplementary Figure.4.** Association of right and left temporal tau ROIs with executive function composite.

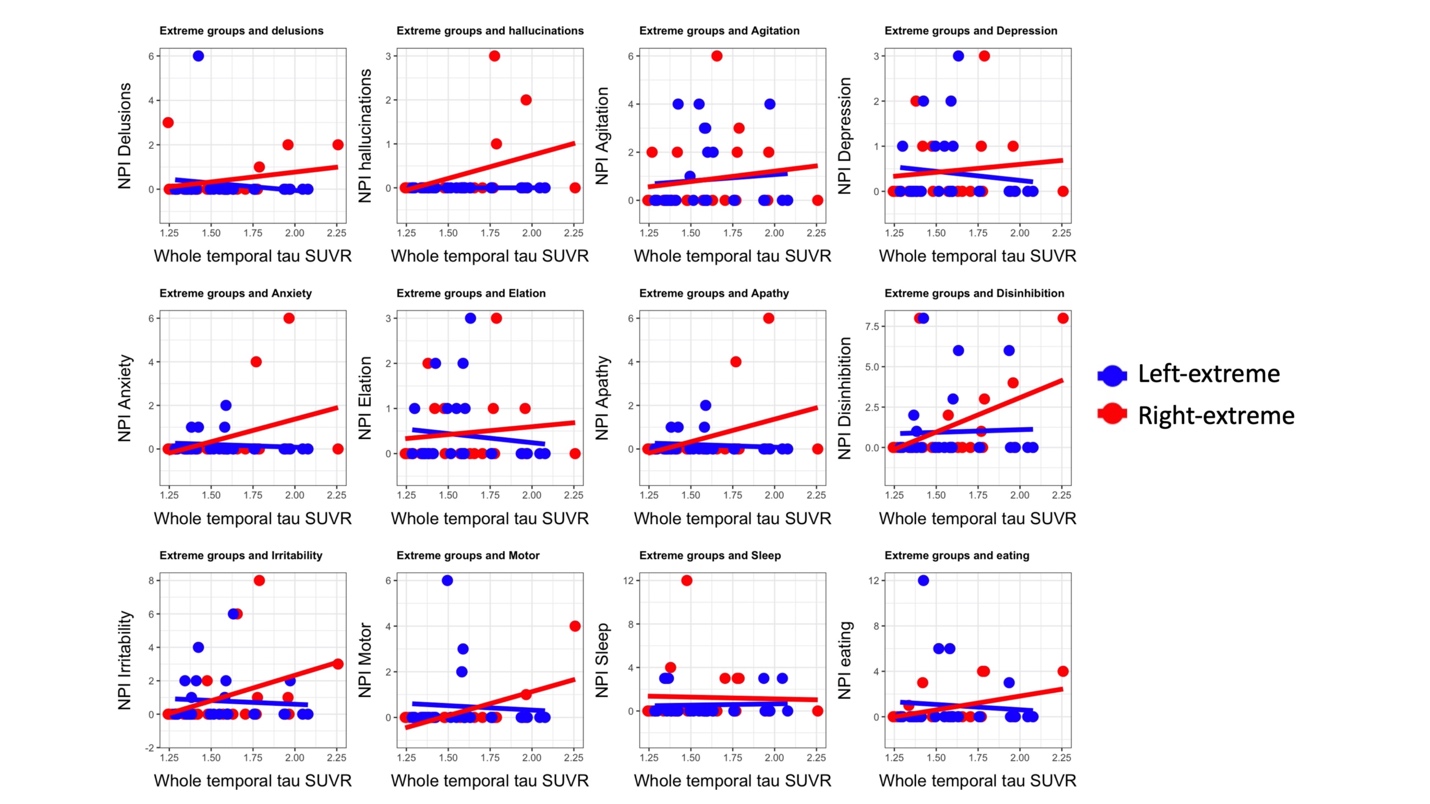

**Supplementary Figure 5.** Association of right and left temporal tau ROIs with neuropsychiatric inventory (NPI) subdomains. Asymmetry-extreme group

| Supplementary Table 1. NPI subdomain in the extreme asymmetry groups | | | | | | |
| --- | --- | --- | --- | --- | --- | --- |
|  | Age | Education | Sex | Group | Whole Temporal Tau | Group x tau |
| A. Delusions | 0.07 (0.2) 0.67 | 0.01 (0.2) 0.94 | -0.35 (0.3) 0.29 | -2.50 (2.0) 0.22 | -0.61 (0.9) 0.52 | 1.63 (1.3) 0.21 |
| **B. Hallucinations** | 0.09 (0.1) 0.22 | 0.12 (0.1) 0.11 | -0.12 (0.1) 0.39 | *-1.64 (0.9) 0.07* | 0.12 (0.4) 0.76 | **1.16 (0.6) 0.04** |
| C. Agitation/Aggression | -0.08 (0.2) 0.7 | 0.08 (0.2) 0.71 | -0.76 (0.5) 0.11 | -1.38 (2.9) 0.63 | 0.18 (1.3) 0.8 | 0.82 (1.8) 0.6 |
| D. Depression/Dysphoria | -0.17 (0.1) 0.22 | -0.09 (0.13) 0.48 | -0.22 (0.3) 0.39 | -1.27 (1.6) 0.42 | -0.73 (0.7) 0.32 | 0.88 (0.9) 0.37 |
| **E. Anxiety** | -0.003 (0.2) 0.9 | 0.19 (0.2) 0.24 | 0.07 (0.3) 0.8 | *-3.72 (2.0) 0.07* | -0.3 (0.9) 0.76 | **2.53 (1.3) 0.05** |
| F. Elation/Euphoria | 0.01 (0.10) 0.93 | 0.08 (0.09) 0.38 | -0.16 (0.2) 0.39 | -1.31 (1.2) 0.27 | -0.48 (0.5) 0.38 | 0.74 (0.7) 0.32 |
| G. Apathy/Indifference | *0.67 (0.37) 0.07* | 0.09 (0.4) 0.79 | -0.38 (0.7) 0.59 | -5.75 (4.3) 0.18 | 1.44 (1.9) 0.47 | 3.57 (2.7) 0.19 |
| H. Disinhibition | 0.13 (0.1) 0.38 | 0.19 (0.14) 0.18 | -0.001 (0.3) 0.99 | -2.02 (1.7) 0.23 | -0.14 (0.8) 0.85 | 1.44 (1.1) 0.18 |
| *I. Irritability/Lability* | -0.03 (0.3) 0.89 | -0.05 (0.28) 0.86 | -0.78 (0.6) 0.16 | *-5.78 (3.4) 0.09* | -0.7 (1.6) 0.67 | *3.78 (2.1) 0.08* |
| J. Aberrant Motor Behavior | 0.28 (0.2) 0.13 | -0.26 (0.17) 0.15 | -0.10 (0.4) 0.76 | -3.43 (2.1) 0.11 | 0.15 (0.9) 0.87 | 1.95 (1.3) 0.15 |
| K. Sleep | 0.06 (0.3) 0.84 | **-0.74 (0.3) 0.02** | 0.64 (0.62) 0.31 | 3.69 (3.8) 0.33 | 0.62 (1.7) 0.72 | -1.74 (2.4) 0.46 |
| L. Appetite and eating disorders | 0.11 (0.4) 0.76 | 0.27 (0.4) 0.47 | 0.13 (0.8) 0.86 | -5.88 (4.6) 0.21 | -0.73 (2.1) 0.72 | 3.52 (2.9) 0.22 |

Bold and shaded green boxes (p<0.05). Italics (p<0.10).

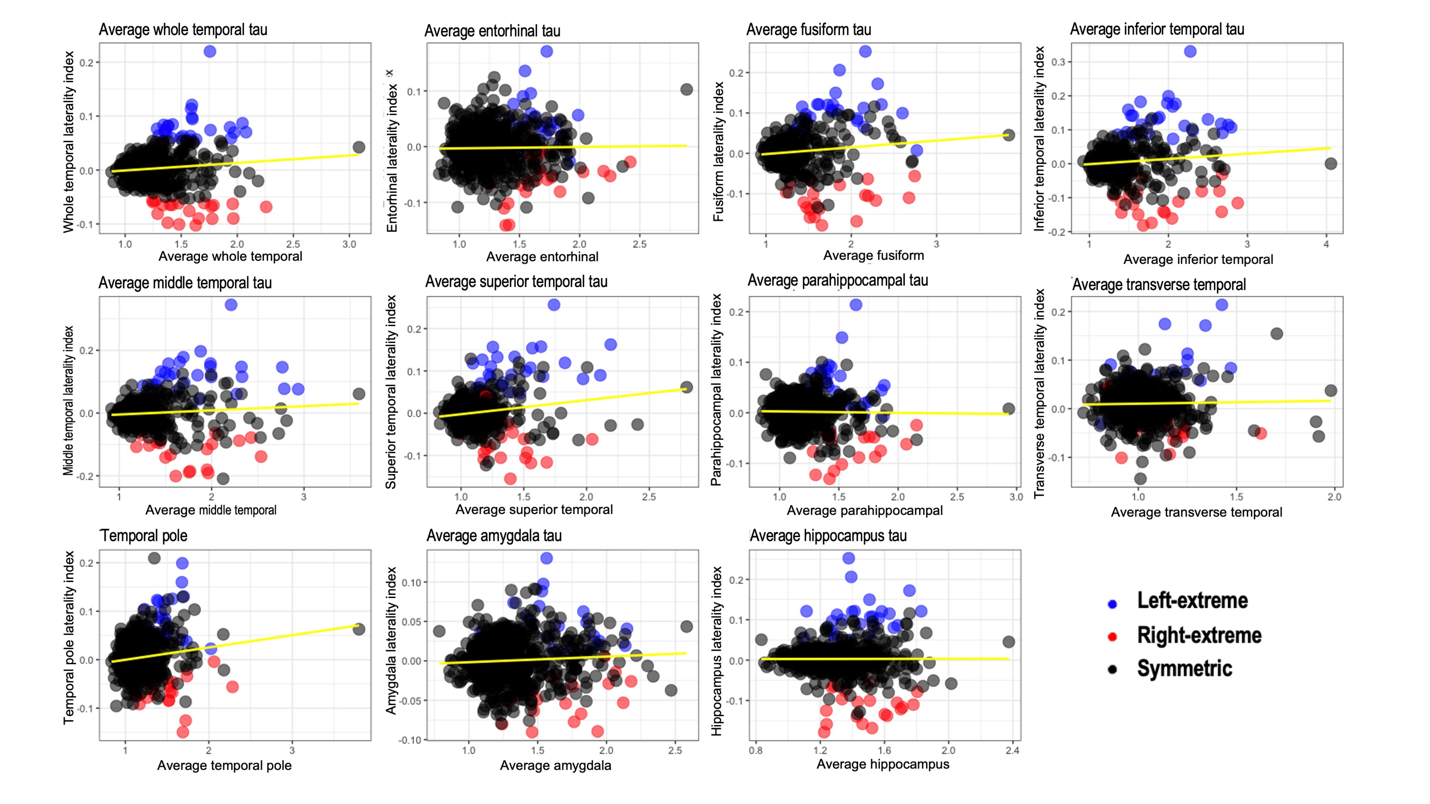

**Supplementary Figure 6.** Correlation of laterality index with tau SUVR

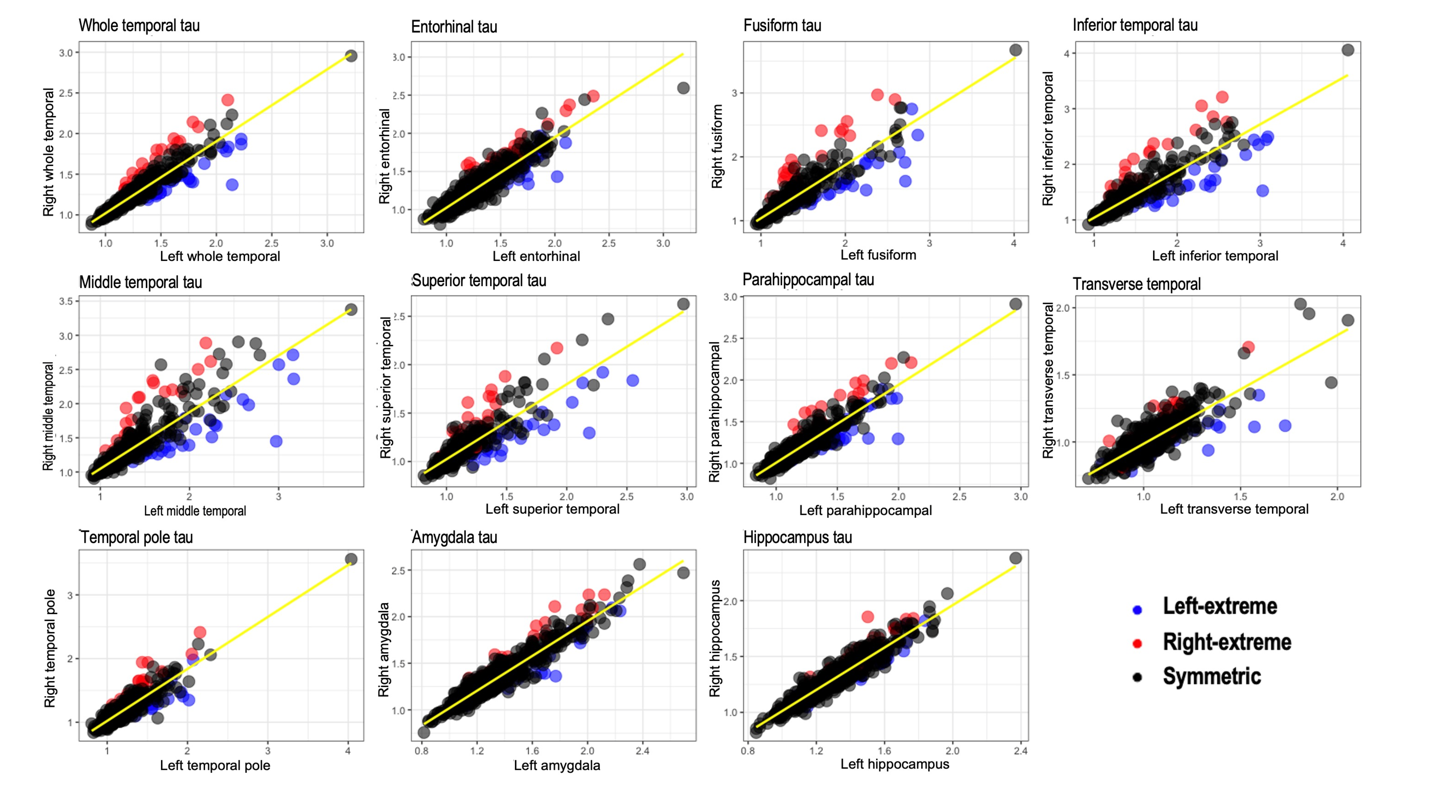

**Supplementary Figure 7.** Correlation of left and right tau ROIs
